# Split ventilation with pressure regulators for patient-specific tidal volumes

**DOI:** 10.1101/2020.07.03.20145409

**Authors:** Lakshminarayan “Ram” Srinivasan, Chris A. Rishel, Barrett J. Larson, Juhwan Yoo, Ned M. Shelton

**Affiliations:** Berkeley, CA; Department of Anesthesiology, Perioperative and Pain Medicine, Stanford University School of Medicine, Stanford, CA; Cupertino, CA; Redwood City, CA

## Abstract

As a measure of last resort during the COVID-19 pandemic, single mechanical ventilators have been repurposed to support multiple patients. In existing split-ventilator configurations using FDA-approved tubing adaptors, each patient receives the same inspiratory pressure, requiring careful matching of patients to avoid barotrauma. Progression of disease may cause tidal volumes to diverge from desired targets, and routine interventions (eg. suctioning) in one patient may adversely affect other patients. To overcome these limitations, we demonstrate a split-ventilator configuration that enables individualized patient management by incorporating a commonly available pressure regulator used for gas appliances. We validate this method by achieving various combinations of tidal volume in each of two synthetic lungs using a standard ventilator machine in combination with two gas flow analyzers. With further safety testing and instrumentation, pressure regulators may represent a viable path to substantially augment the capacity for ventilation in resource-constrained settings.

---

The COVID-19 pandemic has rapidly increased the demand for mechanical ventilators. As a measure of last resort, single mechanical ventilators have been repurposed to support multiple patients. In passive splitting, patients receive identical pressure-control ventilation through a plastic tubing adaptor ^1,2^. This approach has important limitations, recently outlined in a high-profile consensus statement that discouraged the practice^3^. Even after patients are carefully matched, progression of disease may cause tidal volumes to diverge from desired targets, and routine interventions (eg. suctioning) in one patient may adversely affect other patients. In an attempt to overcome these limitations, we set out to develop a split-ventilator configuration that enables individualized patient management by incorporating gas appliance pressure regulators.

Gas appliance pressure regulators are inexpensive and widely available (Figure 1A). They provide natural gas stoves with 18 cm H2O from input line pressures of 35 cmH2O. We sourced two R600S pressure regulators with custom springs (25 oz/in), donated by Maxitrol, Inc., with maximum input pressure of 350 cmH2O, maximum flow rates exceeding 471 mL/min, and adjustable output pressures ranging from 10 to 30 cmH2O. We 3D-printed adapters (3/4” NPT male to 22mm OD) for the regulator input port (ri) and output port (ro). A threaded column in the regulator (1) contains a plastic plunger screw, spring, and cap with gasket to adjust output pressure. An arrow indicates direction of flow; reversing the orientation will result in malfunction. A separate vent on the surface of the regulator should remain open and does not communicate with the patient. We removed the cap during testing, which permitted a small leak, approximately 30 mL per breath.

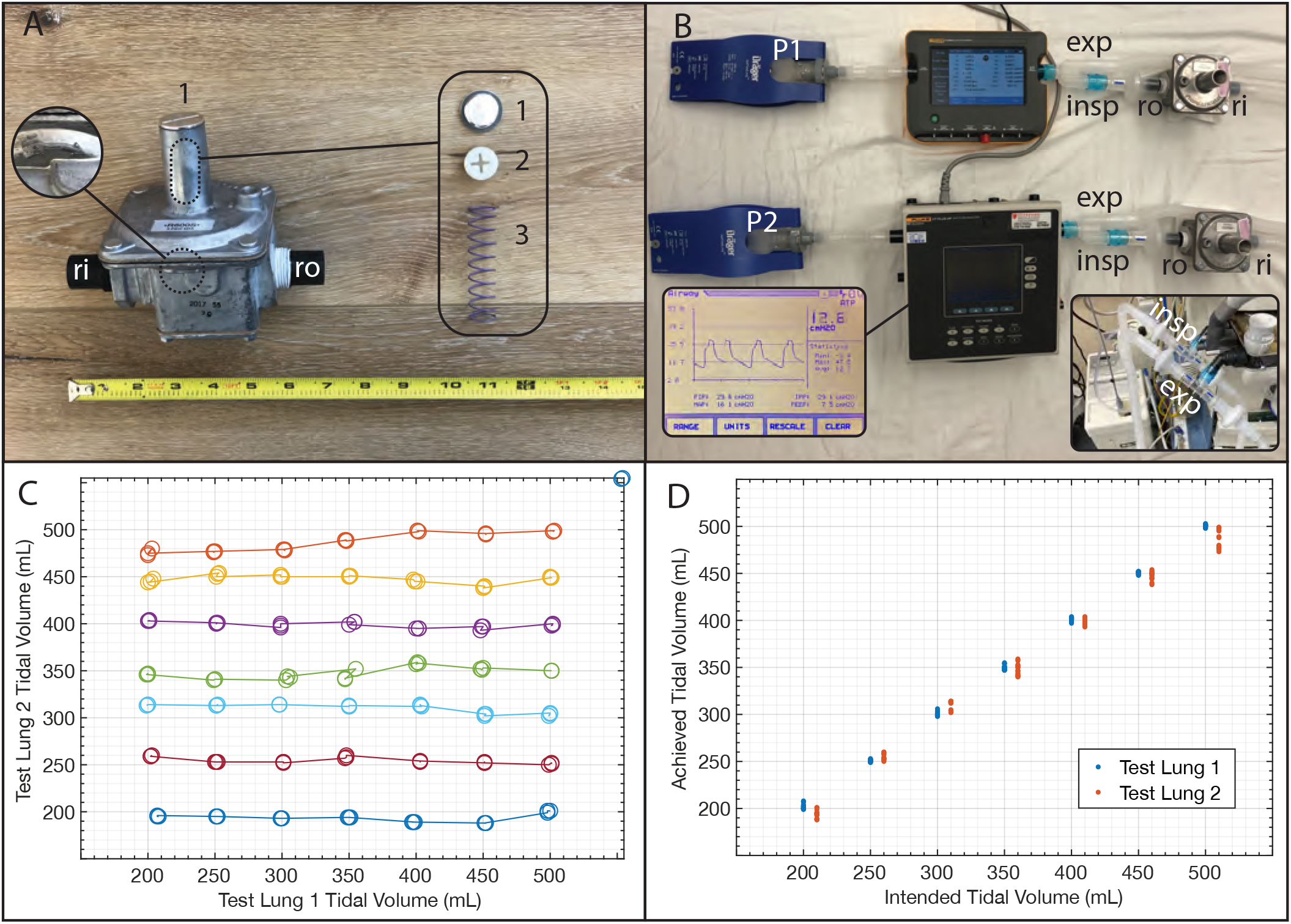
Split ventilation with patient-specific tidal volumes. (A) Gas appliance pressure regulator. (B) Split-ventilator configuration with two gas appliance pressure regulators, one-way valves, and HEPA filters. (C) Various tidal volumes simultaneously achieved in each of two synthetic lungs. (D) Achieved versus intended tidal volume; lung 2 data is rightward-offset by 10 mL on the x-axis for ease of visualization.

To demonstrate feasibility of independent control of patients at two different tidal volumes (TV1, TV2) from a single ventilator, we ventilated two Drager SelfTestLungs (REF MP02400) from a single Drager Apollo anesthesia machine, split using 22mm OD T-pieces and adapters, including HEPA filters and one-way valves to mitigate cross-contamination (Figure 1B). We added one regulator to each inspiratory limb, followed by HEPA filters and one-way valves. The one-way valves also suppressed inter-regulator resonance interactions that would have caused intermittent unregulated pressures at the lowest tidal volume setpoints. Gas analyzers (Fluke VT900A, VT PLUS HF) were placed downstream to each pressure regulator.

By adjusting each regulator separately, we were able to individually control the tidal volumes delivered to each test lung, across a wide range of desired tidal volumes and inter-lung combinations (Figure 1C, 1D). The ventilator was set to pressure-controlled mode with PIP 40 cmH2O, PEEP 8 cmH2O, RR 15, and I:E ratio 1:2. While TV2 was fixed and TV1 was adjusted to achieve 200-500 mL, we quantified no more than 20 mL drift in TV2. Pressures to each test lung varied between 20 to 38 cmH2O.

These data suggest that gas pressure regulators could enable independent tidal volume control during split ventilation, when used with patient-level respiratory monitoring and associated alarms. Further safety testing will be imperative before use in the clinical setting.

## Data Availability

Data is available on request.

## Supplementary Video

“Split ventilation with pressure regulators for patient-specific tidal volumes” L Srinivasan, CA Rishel, BJ Larson, J Yoo, NM Shelton

https://youtu.be/mILLF96YLKo

